# Comparative analysis of SARS-CoV-2 RNA load in wastewater from three different cities of Gujarat, India

**DOI:** 10.1101/2021.04.08.21254861

**Authors:** Vaibhav Srivastava, Shilangi Gupta, Arbind Kumar Patel, Madhvi Joshi, Manish Kumar

**Author notes:** Corresponding Author; **Manish Kumar | Ph.D, FRSC, JSPS, WARI** | Discipline of Earth Science | IIT Gandhinagar | India in. Affiliate | Kiran C Patel Centre for Sustainable Development, IITGN. Member | Global Collaboration on WBE of COVID-19. Member | National Executive Committee, IWA, India. South Asia Representative | METREL Specialist Group, IWA. Editorial Board | Nature Partner Journal (NPJ) Clean Water. Editorial Board | Science of the Total Environment. Editorial Board | Case Studies in Chemical and Environmental Engineering. Editor | Hydrological Research Letter & Groundwater for Sustainable Development. Managing Guest Editor | Journal of Hazardous Material, Elsevier. Managing Guest Editor | Journal of Environmental Management, Elsevier. Google Scholar | Research Gate | ORCID | Twitter | SCOPUS | Impactio* Alt.

## Abstract

The scientific community has widely supported wastewater monitoring of SARS-CoV-2 due to the early and prolonged excretion of coronavirus in the faecal matter. In the present study, eighteen influent wastewater samples from different wastewater treatment plants and pumping stations (5 samples from Vadodara city, 4 from Gandhinagar, and nine from Ahmedabad city) were collected and analyzed for the occurrence of SARS-CoV-2 RNA in Gujarat province, India. The results showed the highest SARS-CoV-2 genome concentration in Vadodara (3078 copies/ L), followed by Ahmedabad (2968 copies/ L) and Gandhinagar (354 copies/ L). The comparison of genome concentration more or less corresponded to the number of confirmed and active cases in all three cities. The study confirms the potential of the Surveillance of Wastewater for Early Epidemic Prediction (SWEEP) that can be used at a large scale around the globe for better dealing with the pandemic situation.

**Graphical abstract:** 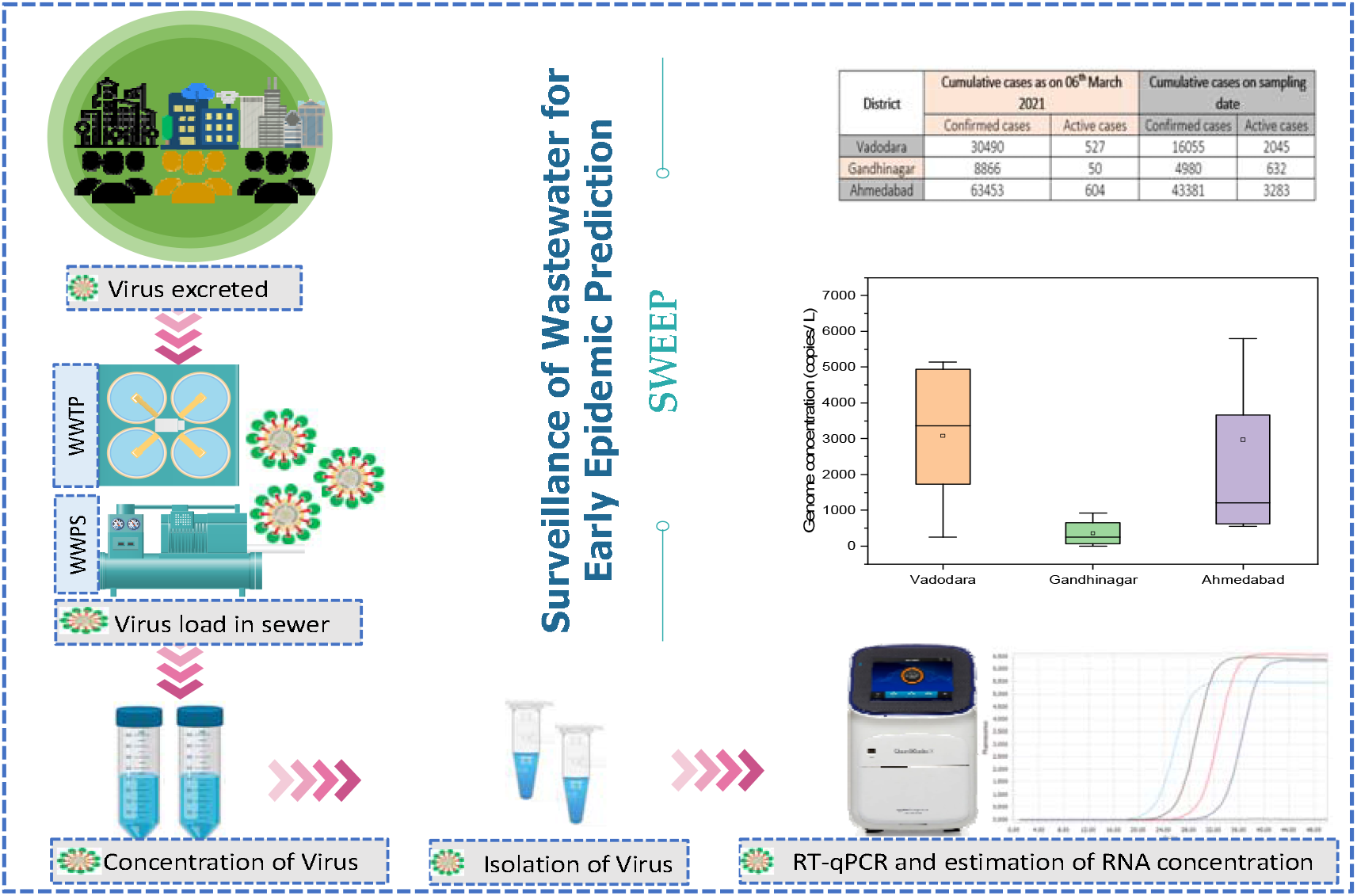

## 1. Introduction

Identifying the emergence and dissemination of the Severe acute respiratory syndrome coronavirus 2 (SARS-CoV-2) biohazard, which remains a global threat after even a year now into the 2019 coronavirus disease (COVID-19) pandemic. While some nations are now attempting to test every person (e.g., Korea and Iceland) to collect population-wide data, this method is inefficient, sluggish, and cost-prohibitive for most nations around the globe [1]. Based on a clinical study, the high pervasiveness of asymptomatic contagious individuals raises doubts about the available data on active cases [2,3]. Wastewater-based epidemiology (WBE) is drawing worldwide attention to COVID-19 surveillance due to the prevalence and protracted exudation of SARS-CoV-2 RNA in the feces of pre-symptomatic and deceased individuals, especially in developed economies with weak health infrastructure.

In India, the first case of COVID-19 was detected when a student returned from Wuhan, China, on 30th January 2020 [4]. Thenceforth, the number of infections has seen a steady spike. India has effectuated international travel bans and a stringent lockdown and curfew in the country to control the spread. Nonetheless, tropical countries like India are at higher risk due to relatively large and dense population demography, inadequate infrastructure, and healthcare services to meet very high demands. Gujarat, India, has recorded 272811 cumulative cases of COVID-19 (active cases: 3025), as of 06^th^ March 2021. The details of the pandemic situation in Vadodara (VABO), Gandhinagar (GN), and Ahmedabad (AMD) have been shown in **Table 1** [5,6,7].

**Table 1.** Comparison of SARS-CoV-2 pandemic in three different cities of Gujarat, India

| District | Cumulative cases as on 06<sup>th</sup> March 2021 |  | Cumulative cases on sampling date |  |
| --- | --- | --- | --- | --- |
|  | Confirmed cases | Active cases | Confirmed cases | Active cases |
| Vadodara | 30490 | 527 | 16055 | 2045 |
| Gandhinagar | 8866 | 50 | 4980 | 632 |
| Ahmedabad | 63453 | 604 | 43381 | 3283 |

To better understand the skill and possible implementation of WBE surveillance of the novel coronavirus, the wastewater analysis for the occurrence of SARS-CoV-2 RNA was performed in three different cities (VABO, GN, and AMD) Gujarat, India, and comparisons were made with the clinical survey-based data. We also studied the temporal variance in the viral RNA concentration in STPs during post lockdown time in GN and AMD cities of Gujarat, India [8,9]. The prime goal of the present study was to substantiate the Surveillance of Wastewater for Early Epidemic Prediction (SWEEP) potential to know the extent of COVID infection by comparing the SWEEP data with clinical survey-based secondary data. Also, it will persuade the authorities and policymakers to incorporate WBE surveillance into the regular monitoring program and policy framework to manage current or future COVID-19 like pandemic situations efficiently.

## 2. Materials and methods

### 2.1 Sampling location

In the present study, eighteen influent wastewater samples (5 samples from Vadodara city, 4 from Gandhinagar, and nine from Ahmedabad city) were collected and analyzed for the presence of SARS-CoV-2 genetic material **(Fig. 1)**. In all three cities, the sewage is collected through a system comprising an underground drainage network, auxiliary pumping stations (APS), Sewage Treatment Plant, and disposal into the natural water bodies and rivers after treatment. Wastewater generated from all this development is collected by a network of underground sewers and pumping stations and is conveyed to the sewage treatment works for physical and biological treatment to meet the Gujarat Pollution Control Board (GPCB) guidelines before discharge into the nearest water body.

**Fig. 1.**
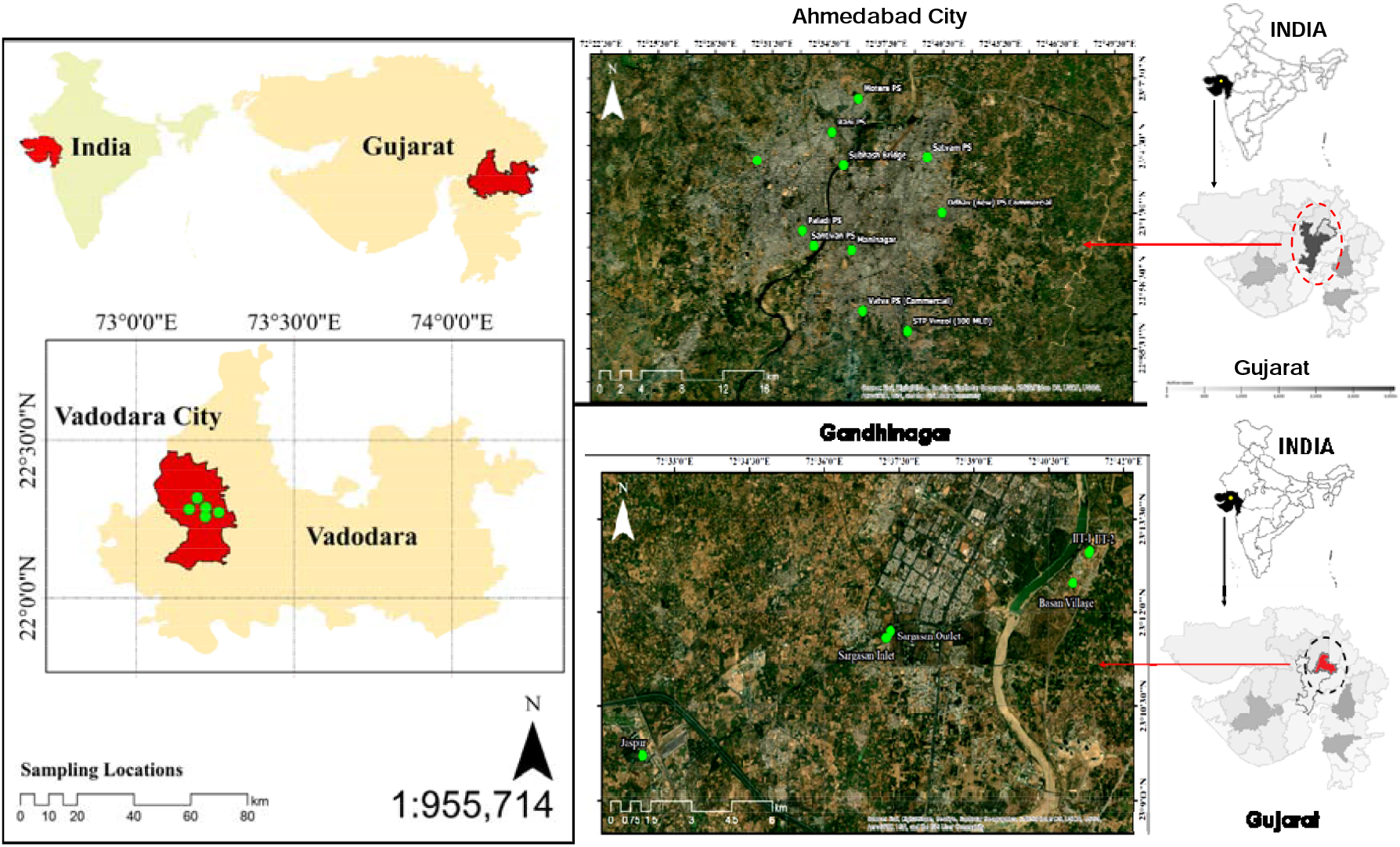
Geospatial position of sampling locations in three different cities of Gujarat, India

Vadodara Municipal Corporation has three drainage zones for the sewerage system based on the city’s natural topography. Each of the drainage zones has a sewage treatment plant (STP). The sewage from drainage zones-I and II is disposed into the Jambuva River which, ultimately joins the River Vishwamitri. The sewage from drainage zone-III is disposed into the River Vishwamitri. The schematic layout of the wastewater treatment in Vadodara is shown in **Fig. 2**. Sewage Disposal Works Department of Vadodara includes 6 STPs & 49 Auxiliary/Main Pumping stations (APS/MPS). In the APS, the wastewater (sewage) from various parts of the city is collected in the wet well of the APS and then pumped to the Main Pumping Station and ultimately to the STP for treatment. Based on the natural topography of the Vadodara city sewerage system is divided into three drainage zones.

**Fig.2.**
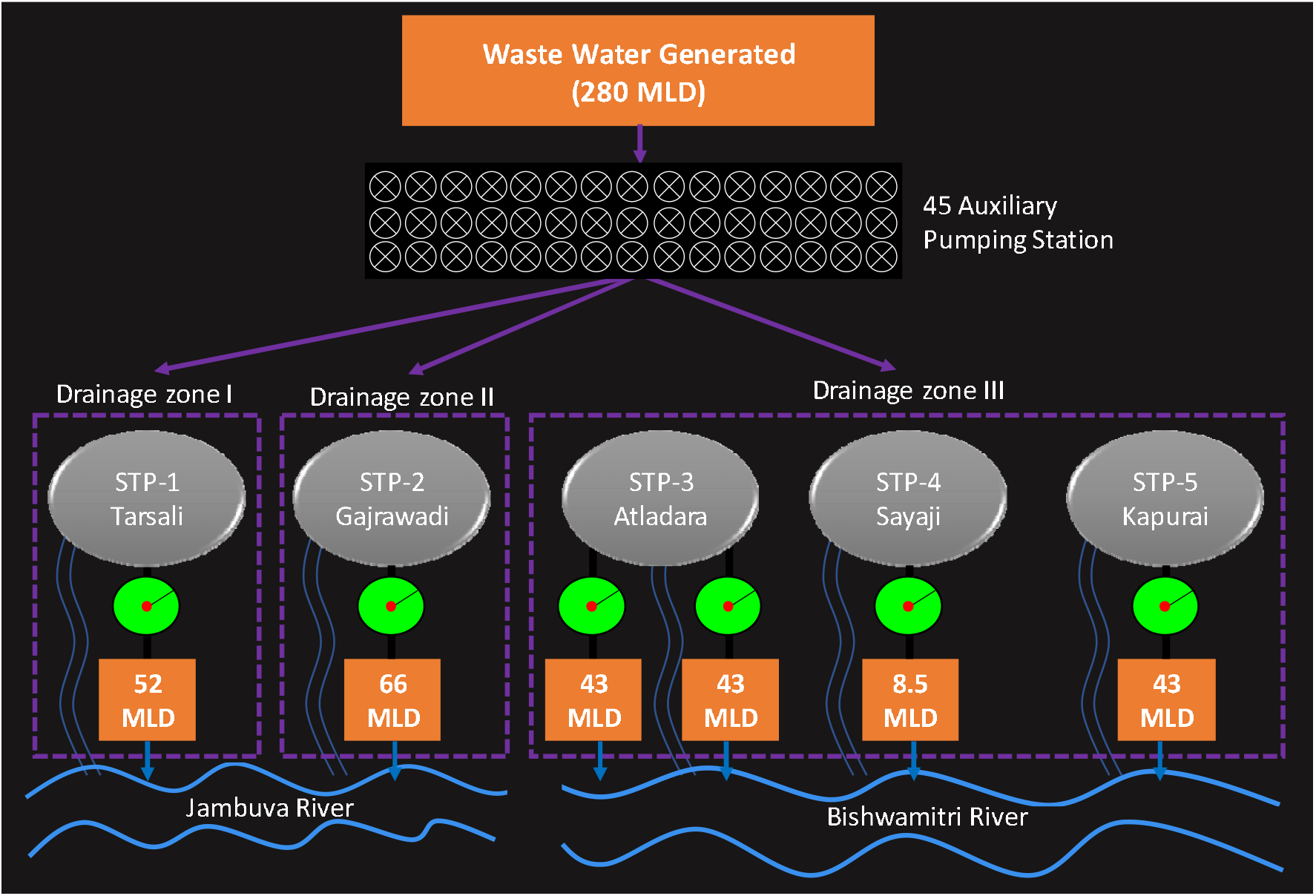
Sampling points and layout of the wastewater treatment in Vadodara, Gujarat, India

Likewise, the Ahmedabad Municipal Corporation comprises 9 STPs, 45 Sewage Pumping Stations, and an extended Sewage Network of ∼2500 km present in the city. In Gandhinagar, the entire city’s wastewater is first collected in the Sargasan Drainage Pumping Station via the underground pipe network. Thereafter, it is pumped and transferred mainly to the Jaspur and Sargasan STPs, where treatment processes occur. Details of the sampling locations, such as geospatial positions, capacity of the treatment plant, and wastewater source, are given in **Table 2**.

**Table 2.**
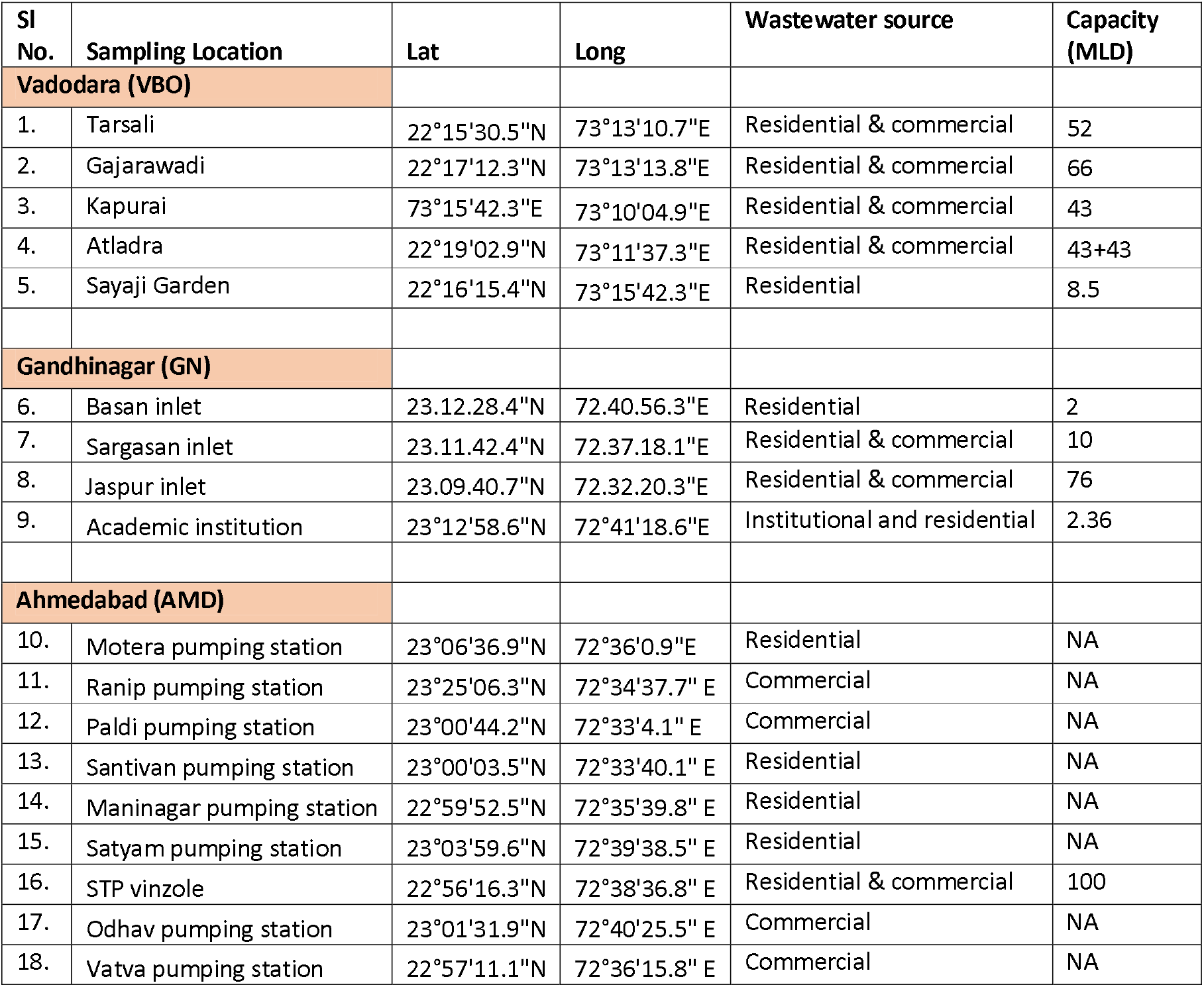
Details of Sampling locations

### 2.2 Sample collection and preparation

The untreated wastewater was collected from different locations in three cities, i.e., Vadodara (VABO), Gandhinagar (GN), and Ahmedabad (AMD) of Gujarat province, India. A total of 5 influent samples were collected from five STPs of VABO, 4 samples from STPs in GN, and 9 samples (8 from pumping stations and one from STP) from AMD in the first week of November 2020. The grab sampling method was used for the sample collection in 500ml polyethylene sterile bottles (Tarsons, PP Autoclavable, Wide Mouth Bottle, Cat No. 582240, India). Collected samples were transferred in an icebox to the laboratory and refrigerated at 4^0^ C until further process. A sampling blank was also prepared to examine the cross-contamination during transportation. The experiments were performed at Gujarat Biotechnology Research Centre (GBRC), an approved laboratory by the Indian Council of Medical Research (ICMR), New Delhi.

### 2.3 Concentration methods

The concentration method consisted of a PEG 9000 (80 g/L) and NaCl (17.5 g/L) precipitation protocol previously described by Kumar et al., 2020 [10] for wastewater samples. 30ml sample was centrifuged (Model: Sorvall ST 40R, Thermo Scientific) at 4000g for 30 minutes in a 50ml falcon tube followed by the filtration of the supernatant with a syringe filter of 0.2µ (Mixed cellulose esters syringe filter, Himedia). The 25ml sample filtrated was then treated with the NaCl (17.5 g/L) and PEG 9000 (80 g/L) and incubated at 17°C, 100 rpm overnight (Model: Incu-Shaker™ 10LR, Benchmark). The sample was then transferred in an oak ridge tube for further centrifugation (Model: Incu-Shaker™ 10LR, Benchmark) at 14000g for 90 minutes, ultimately forming the pellets. RNase-free water was used for the resuspension of the viral particles after discarding the supernatant. The sample was then stored in a 1.5ml Eppendorf tube at a temperature of −40°C for RNA isolation.

### 2.4 Isolation of the SARS-CoV-2 viral genome

Using a commercially ready-for-use kit (NucleoSpin^®^ RNA Virus, Macherey-Nagel GmbH & Co. KG, Germany), SARS-CoV-2 RNA isolation was performed. MS2 phage (10 μL), Proteinase K (20 μL) and RAV1 buffer (600 μL) consisting of carrier RNA were mixed with 300 μL of the concentrated viral particles. MS2 phage serves as the molecular process inhibition as a test control [11]. It was used to monitor the efficacy of RNA extraction and PCR inhibition. It should be remembered that MS2 may spontaneously exist in wastewater, so there is a risk that the retrieved MS2 may consist of both the spiked and the background viral material. As per the user manual instructions (Macherey-Nagel GmbH & Co. KG), further procedures were carried out. The last elution was done with 30 μL of kit-supplied elution buffer. Using a Qubit 4 Fluorometer (Invitrogen), RNA concentrations were checked.

The nucleic acid was analyzed to identify the S gene, N gene, and ORF1ab of SARS-CoV-2 and the internal control (MS2) with the help of RT-PCR using the TaqPath™ Covid-19 RT-PCR package (Applied Biosystems). Amplification was conducted in a reaction (25 μL) vial containing 7 μL of RNAs derived from each sample. 2 μL of the positive control (TaqPath™ COVID-19 Control) and refined 5 μL of negative control were used for the study. Nuclease-free water was applied as a template-free control in this analysis. Additional process steps were executed, as defined in the product guidebook. The RT-qPCR step consisting of 40 cycles, included UNG incubation (25 °C for 2 min), reverse transcription (53 °C for 10 min), and activation (95 °C for 2 min). The reactions were conducted and elucidated as instructed in the handbook of Applied BiosystemsTM 7500 Fast Real-Time PCR.

### 2.5 Data visualization

OriginPro 2019b software has been used for data analysis and to draw boxplots.

## 3. Results and discussion

Wastewater samples collected from three cities (Vadodara, Gandhinagar, and Ahmedabad) of Gujarat, India, showed a great variation in SARS-CoV-2 RNA load. Comparison of RT-PCR assay findings for the detection of SARS-CoV-2 RNA (N, ORF 1ab, and S genes) among three cities showed 100% positive samples in Vadodara (5/5), 75% in Gandhinagar (3/4), and 100% in Ahmedabad (9/9). The average N-gene copies were found to be maximum in AMD (4731 copies/L), followed by VABO (3179 copies/ L) and GN (243 copies/ L). The ORF 1ab-gene copies were found maximum in wastewater samples collected from VABO (3730 copies/ L), followed by AMD (2756 copies/ L) and GN (611 copies/ L). Similarly, the descending order of S-gene copies was: VABO (2325 copies/ L)> AMD (1417 copies/ L)> GN (207 copies/ L). Conclusively, a greater genome concentration was noticed in VABO (3078 copies/ L), trailed by AMD (2968 copies/ L) and GN (354 copies/ L). The distribution of SARS-CoV-2 gene copies in wastewater samples collected from three cities is depicted in **Fig. 3**. Also, the variation in gene copies of the SARS-CoV-2 targeted genes and genome concentration in wastewater samples is shown in **Table 3**.

**Table 3.** Variation in gene copies of the SARS-CoV-2 targeted genes and genome concentration in wastewater samples, collected from three different cities of Gujarat, India

| Sampling date | Station | Ct value |  |  | Gene copies (copies/ L) |  |  |  |
| --- | --- | --- | --- | --- | --- | --- | --- | --- |
|  |  | N | ORF | S | N | ORF | S | Genome concentration |
| 02.11.2020 | **Vadodara** |  |  |  |  |  |  |  |
|  | STP-1 Tarsali | 29.9 | 30.31 | 31.43 | 4814 | 3594 | 1656 | 3355 |
|  | STP-2 Gajrawad | 31.09 | 31.1 | 32.16 | 2086 | 2069 | 1012 | 1722 |
|  | STP-3 Atladara | 30.04 | 29.53 | 29.91 | 4346 | 6261 | 4792 | 5133 |
|  | STP-4 Sayaji | 33.76 | 34.18 | 35.96 | 360 | 275 | 93 | 243 |
|  | STP-5 Kapurai | 30.06 | 29.49 | 30.13 | 4292 | 6449 | 4072 | 4938 |
| 02.11.2020 | **Gandhinagar** |  |  |  |  |  |  |  |
|  | Basan Inlet | 32.65 | 31.53 | 33.32 | 733 | 1551 | 474 | 919 |
|  | Jaspur Inlet | 36.00 | 34.53 | 36.91 | 91 | 222 | 53 | 122 |
|  | IIT inlet | ND | ND | ND | 0 | 0 | 0 | 0 |
|  | Sargasan inlet | 35.20 | 32.78 | 34.04 | 147 | 672 | 301 | 373 |
| 05.11.2020 | **Ahmedabad** |  |  |  |  |  |  |  |
|  | Motera PS | 30.63 | 33.65 | 33.73 | 2873 | 386 | 365 | 1208 |
|  | Ranip PS | 29.50 | 30.58 | 31.50 | 6415 | 2986 | 1580 | 3661 |
|  | Paldi PS | 32.39 | 33.02 | 33.79 | 869 | 577 | 352 | 599 |
|  | Santivan PS | 31.57 | 31.88 | 32.95 | 1506 | 1220 | 603 | 1110 |
|  | Maninagar PS | 32.46 | 32.98 | 34.57 | 830 | 593 | 217 | 547 |
|  | Satyam PS | 32.14 | 32.61 | 36.50 | 1024 | 752 | 68 | 615 |
|  | STP Vinzole | 28.75 | 30.03 | 31.27 | 11148 | 4386 | 1848 | 5794 |
|  | Odhav PS | 28.31 | 28.52 | 29.39 | 15523 | 13247 | 6930 | 11900 |
|  | Vatva PS | 30.90 | 32.83 | 32.53 | 2390 | 654 | 793 | 1279 |

**Fig.3.**
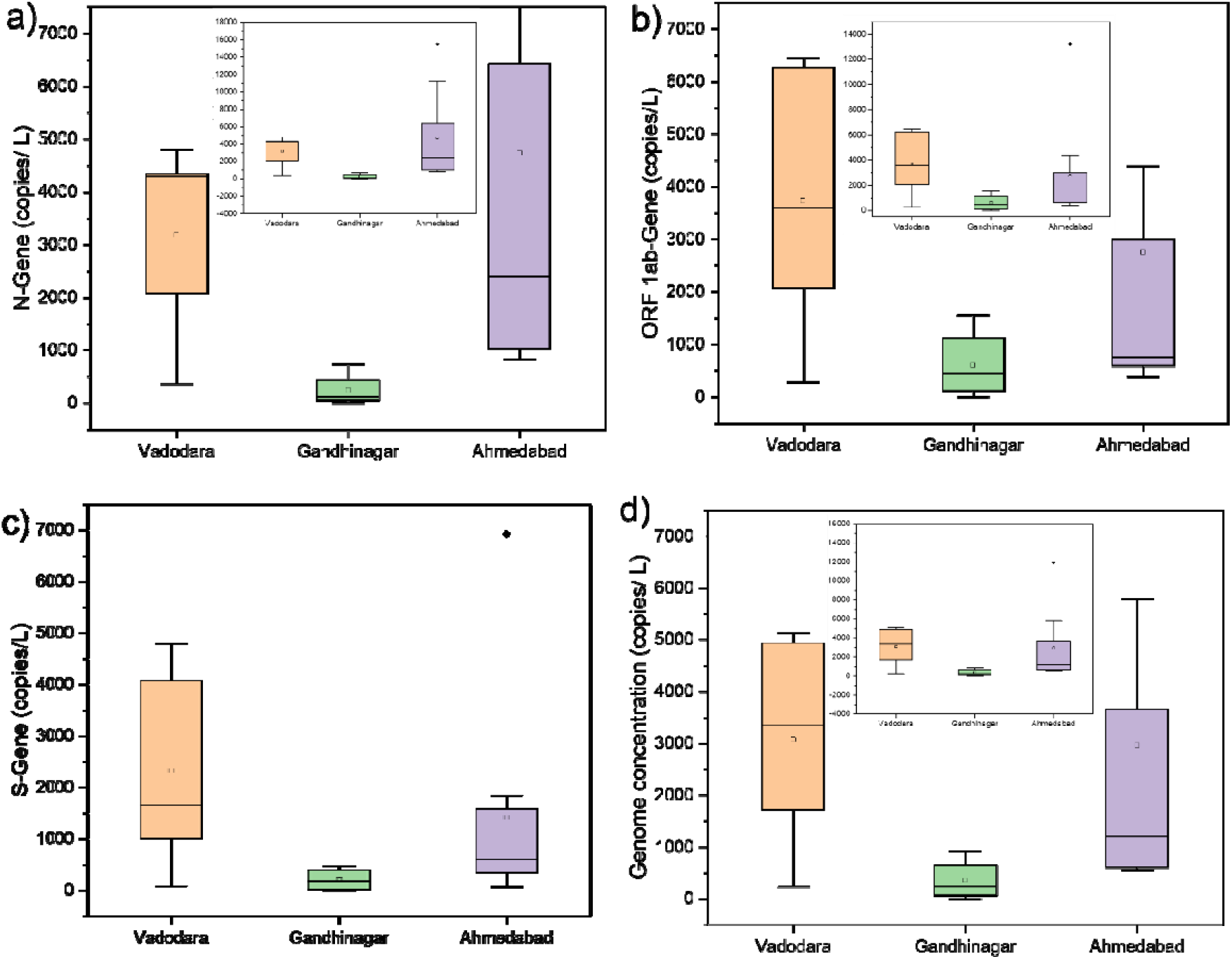
Distribution of SARS-CoV-2 gene copies, collected from three different cities of Gujarat, India; a.) N gene, b.) ORF 1ab gene, c.) S gene, and d) Genome concentration

The trends of virus genetic load were more or less in line with the number of confirmed and active cases, which were highest in VABO, followed by AMD and GN **(Table 1)**. A very nominal difference in the SARS-CoV-2 genome concentration in wastewater samples of VBO and AMD was noticed despite a difference of more than two-folds in the cumulative number of confirmed cases and above 1000 active cases in AMD compared to VBO on sampling date **(Table 1)**. This trend can be ascribed to the fact that samples were collected from STPs (5) in VABO; while in the case of AMD, samples were mainly collected from pumping stations (8). Therefore, the concentration of SARS-CoV-2 RNA might be higher in STPs as compared to the sewage pumping station that was reflected in the analysis in VABO. However, some other factors such as population density, city development plan, sewerage system, health amenities, and management strategies may influence the SARS-CoV-2 genetic load in wastewater samples.

The results were in agreement with Kumar et al. [8], who studied weekly temporal variation in SARS-CoV-2 genetic material concentration in wastewater samples targeting N, ORF 1ab, and S genes in a two-month study in Gandhinagar. The results suggested a positive correlation between SARS-CoV-2 genome concentration in wastewater and the number of confirmed cases, which was found higher in the month of September compared to August 2020, corresponded to ∼2.2 folds increase in confirmed cases during the study period. Likewise, in another three-month (September to November 2020) weekly analysis of wastewater samples from 9 different locations in Ahmedabad city showed similar trends, and the maximum SARS-CoV-2 genome concentration was noticed in November (∼10729 copies/ L), trailed by September (∼3047 copies/ L), and October (454 copies/ L) in line with a ∼ 1.5-fold rise in the confirmed cases during the study period. The decrease in SARS-CoV-2 concentration in October subjected to a decline of 20.5% in active cases (∼844 cases), while a significant rise in virus RNA in November 2020 was due to a rise of 1.82% in active cases (∼59 cases). Though the rise in active cases was nominal in November, but at the same time, a sharp rise of >7000 new cases (17.3%) was reported in November 2020 [9]. There are many other studies in the public domain from different parts of the world, such as the Netherlands [3], Spain [12], the USA [13,14], Paris [15], China [16], India [8,10], Australia [17], etc. which support WBE surveillance of COVID-19.

One of the main advantages of WBE is that it includes both asymptomatic and symptomatic individuals, therefore can give a better picture of the pandemic situation as compared to clinical-based secondary data, which includes only symptomatic patients and rely on the number and efficiency of clinical tests. Therefore, under certain circumstances, it is possible that despite an increase in SARS-CoV-2 RNA load in wastewater, no significant change in COVID cases may observe. Consequently, SWEEP technology can provide the actual extent of the infection at sub-city or zone levels and help in identifying the hot spots within a city.

## 4. Conclusion

A comparison of SARS-CoV-2 RNA presence in wastewater samples from three cities of Gujarat unveiled the highest load in VBO, followed by AMD and GN. The virus genetic material showed a positive correlation with the number of confirmed and active cases in all three cities. Also, the genome concentration more or less corresponded to the number of confirmed and active cases in the present study. The study concludes that regular monitoring of wastewater samples could be used to know the pandemic situation in a particular area and help in tuning the management interventions efficiently. Though WBE has immense potential that must be exploited and included in the policy framework around the globe; however long-scale time-series data along with epidemiological information is required to substantiate the robustness of this technology. Also, future emphasis should be paid to developing a predictive model using WBE and clinical survey data for a better understanding of the situation to the policymakers and enhancing the preparedness and management of epidemic/ pandemic situations.

## Notes

The authors declare no competing financial interest.

## Data Availability

All data is included in the paper.

## Acknowledgement

This work is funded by UNICEF, Gujarat and UKIERI. We also acknowledge the help received from Mr. Alok Thakur, and other GBRC staffs who contributed towards sample and data analyses.

## References

[1] Hart, O E., Halden, R. U. 2020. Computational analysis of SARS-CoV-2/COVID-19 surveillance by wastewater-based epidemiology locally and globally: Feasibility, economy, opportunities and challenges. Sci. Total Environ. 730, 138875. https://doi.org/10.1016/j.scitotenv.2020.138875

[2] Rimoldi, S. G., Stefani, F., Gigantiello, A., Polesello, S., Comandatore, F., Mileto, D., Maresca, M., Longobardi, C., Mancon, A., Romeri, F., Pagani, C., Moja, L., Gismondo, M.R., Salerno, F.,2020. Presence and vitality of SARS-CoV-2 virus in wastewaters and rivers. Sci. Total Environ. 744:140911

[3] Medema, G., Heijnen, L., Elsinga, G., Italiaander, R., Brouwer, A., 2020. Presence of SARS Coronavirus-2 RNA in sewage and correlation with reported COVID-19 prevalence in the early stage of the epidemic in the Netherlands. Environ Sci Technol Lett. 7 (7), 511–516. https://doi.org/10.1021/acs.estlett.0c00357

[4] WHO. Coronavirus disease (COVID-19), India situation report. WHO India 2020. https://www.who.int/india/emergencies/india-situation-report

[5] Amdavad Municipal Corporation, Gujarat, India. 2021. https://ahmedabadcity.gov.in/portal/web?requestType=ApplicationRH&actionVal=loadCoronaRelatedDtls&queryType=Select&screenId=114

[6] Gandhinagar Municipal Corporation, Gujarat, India. 2021. http://gandhinagarmunicipal.com/corona_dashboard/

[7] COVID India, Gujarat District Wise COVID Data, Gujarat, India. 2021. https://www.covid19india.org/state/GJ

[8] Kumar, M., Joshi, M., Patel, A. K., & Joshi, C. G., 2021a. Unravelling the early warning capability of wastewater surveillance for COVID-19: A temporal study on SARS-CoV-2 RNA detection and need for the escalation. Environ. Res. 110946. https://doi.org/10.1016/j.envres.2021.110946

[9] Kumar, M., Joshi, M., Shah, A. V., Srivastava, V., & Dave, S., 2021b. First wastewater surveillance-based city zonation for effective COVID-19 pandemic preparedness powered by early warning: A study of Ahmedabad, India. medRxiv. https://doi.org/10.1101/2021.03.18.21253898

[10] Kumar, M., Patel, A.K., Shah, A.V., Raval, J., Rajpara, N., Joshi, M. and Joshi, C.G., 2020. First proof of the capability of wastewater surveillance for COVID-19 in India through detection of genetic material of SARS-CoV-2. Sci. Total Environ. 746, p.141326. https://doi.org/10.1016/j.sci378totenv.2020.141326

[11] Haramoto, E., Kitajima, M., Hata, A., Torrey, J.R., Masago, Y., Sano, D., Katayama, H., 2018. A review on recent progress in the detection methods and prevalence of human enteric viruses in water. Water Res. 135, 168–186. https://doi.org/10.1016/j.watres.2018.02.004

[12] Randazzo, W., Truchado, P., Cuevas-Ferrando, E., Simón, P., Allende, A., Sánchez, G., 2020. SARS-CoV-2 RNA in wastewater anticipated COVID-19 occurrence in a low prevalence area. Water Res. 526 (1), 135–140. https://doi.org/10.1016/j.bbrc.2020.03.044181, 115942.

[13] Wu, Y., Guo, C., Tang, L., Hong, Z., et al., 2020a. Prolonged presence of SARS-CoV-2 viral RNA in faecal samples. Lancet Gastroenterol. Hepatol. 5, 434–435. https://doi.org/10.1016/S2468-1253(20)30083-2.

[14] Nemudryi, A., Nemudraia, A., Surya, K., Wiegand, T., Buyukyoruk, M., Wilkinson, R., Wiedenheft, B., 2020. Temporal detection and phylogenetic assessment of SARSCoV-2 in municipal wastewater. medRxiv https://doi.org/10.1101/2020.04.15.200667462020.03.22.20041079.

[15] Wurtzer, S., Marechal, V., Mouchel, J.M., Moulin, L., 2020. Time course quantitative detection of SARS-CoV-2 in Parisian wastewaters correlates with COVID-19 confirmed cases. medRxiv https://doi.org/10.1101/2020.04.12.200626792020.04.12.20062679

[16] Zhang, W., Du, R.H., Li, B., Zheng, X.S., Yang, X. Lou, Hu, B., Wang, Y.Y., Xiao, G.F., Yan, B., Shi, Z.L., Zhou, P., 2020. Molecular and serological investigation of 2019-nCoV infected patients: implication of multiple shedding routes. Emerg. Microbes Infect. 9 (1), 386–389. https://doi.org/10.1080/22221751.2020.1729071.

[17] Ahmed, W., Angel, N., Edson, J., Bibby, K., Bivins, A., O’Brien, J. W., & Mueller, J. F., 2020. First confirmed detection of SARS-CoV-2 in untreated wastewater in Australia: a proof of concept for the wastewater surveillance of COVID-19 in the community. Sci. Total Environ. 728, 138764. https://doi.org/10.1016/j.scitotenv.2020.138764

